# *I had to leave. I had to leave my clinic, my city, leave everything behind in Syria*. Qualitative accounts from Syrian health care workers migrating from the war-torn country

**DOI:** 10.1101/2020.09.19.20178103

**Authors:** Agneta Kallström, Al Abdulla Orwa, Parkki Jan, Mikko Häkkinen, Hannu Juusola, Kauhanen Jussi

## Abstract

**Objectives:** To explore the experiences of Syrian health care workers of violence since 2011 and reasons behind migration from the conflict-affected country

**Design:** A qualitative study using semi-structured interviews and inductive content analysis. Interviews were conducted in Turkey and Europe in 2016 - 2017.

**Setting:** Interviewees were Syrian health care workers who had been working in the country after the conflict started in 2011, but at some point, migrated from Syria to Turkey or Europe.

**Participants:** We studied data from 20 semi-structured in-depth interviews collected with a snowball sampling method.

**Results:** Our findings show that health care workers migrated from Syria only because of security concerns. In most cases, the decision to leave was a result of the generalised violence by different warring parties. Targeted attacks against health care was one of the main reasons for leaving. Some participants had a specific notable trigger event before they left; such as a colleague being detained or killed. Many just grew tired of living under constant threat and fear, with their families also at risk.

**Conclusions:** This research adds to the body of literature on violence in Syria. It helps to understand the reasons why health care workers, even though realising that it will leave their population without proper health care provision, nevertheless decide to flee the country. Understanding the motives of the health care workers will help to find new ways to protect them.

## INTRODUCTION

Armed conflicts challenge regular health care provision. Health care workers (HCWs), one of the most crucial factors of health care services, often migrate away from conflict-affected areas.[1–3] In 2019, at least 51 HCWs in 17 countries were killed and 502 injured.[4]Notably, attacks against health care occurred during the events known as the Arab Spring, a revolutionary wave of demonstrations and protests starting in 2010 from Tunisia. Anti-government demonstrations escalated into violence in several states around the Middle East and brought down regimes. In Syria, the events turned into an armed conflict that is still ongoing. Especially in Syria, Bahrain, Yemen and Iraq, the HCWs is targeted since the start of the Arab Spring.[5–7]

During the decade of war, hospital bombings have become a trademark in the Syrian conflict. Violence against health care and thus denying the provision of health care to the Syrian population has been used as a strategy of war[7] even though International humanitarian law (IHL) stipulates that the health care system is protected in time of war. The government must protect health care workers’ medical neutrality, which means freedom for the HCWs to take care of patients regardless of their political affiliations[8]. From the beginning of the conflict, the Government of Syria (GoS) is reported to punish HCWs for treating injured protestors[9,10] regardless of the IHL statute that *under no circumstances shall any person be punished for carrying out medical activities compatible with medical ethics, regardless of the person benefiting from them*.[11] In 2012 the GoS effectively criminalised health care provision to people belonging to the opposition.[10,12]

At least 923 HCWs have been killed in Syria from 2011 through March 2020. All warring parties have attacked health care; however, the GoS with Russia has been held responsible for 91 per cent of deaths of HCWs. Non-state armed groups (NSAGs), Islamic State in Iraq and Syria (ISIS), Kurdish or unidentified forces are responsible for 9 per cent. The primary cause of death (55%) is aerial or ground bombardment. HCWs have also died when detained and tortured, or just executed.[4,13,14] More than 70 % of qualified HCWs have left the country, in some areas, specific specialities are totally absent.[15,16] Also, medical students have had to leave their basic and specialisation studies when fleeing violence.[17] The absence of professionals will add to the challenges of both current and future treatment of the population.

HCWs, especially physicians, tend to migrate at the early stage of the conflict because they have resources to leave.[18] The financial issues and training concerns predict HCWs migration. Also, generalised insecurity, targeted violence and protecting family is a known reason to go.[19,20] In Iraq, a country that has experienced perpetual violence for decades, male physicians over 30 years of age face a significantly increased risk of being kidnapped or assassinated. However, factors demoralising the HCWs are complex, and the decision to leave is made based on a sum of reasons. [1,21]

Most analyses of violence against health care in Syria have focused on damage to health care infrastructures, such as hospitals and ambulances.[22–24] Few studies have looked at HCWs personal experiences in conflicts[15,25], and none of them examines the reasons for HCWs leaving specifically Syria. Also, current reviews do not take into account how the violence by different warring parties, such as arrests, detention and kidnappings, affect the decision to leave to protect their own life and family members. Mainly missing is data on secondary trauma, such as witnessing killings of civilians and colleagues, as well as how the constant fear of the conflict itself affects the outward mobility of health care professionals.

Many studies focus on the staff departing less developed countries for economic reasons, or due to inadequate quality health care.[21,26] Still, there is a relative lack of studies on healthcare-related migration in conflict settings.

Doocy et al. [27] showed that fleeing from Iraq was associated with a violent event in 61 per cent of cases. The physicians who left the country, a total of 75 per cent had experienced violence against their household before the decision to leave. In another study by Al-Khalisi [28], 60 per cent of participants had left Iraq for security reasons.

Violence in Iraq has differed from that in Syria with almost no airstrikes against health care facilities there. On the contrary, Fouad et al. [7] have argued that the bombing of hospitals is part of the weaponisation of health care, a strategy of war, by the Government of Syria (GoS).

## METHODS

In this research, we studied HCWs who have migrated from Syria and reside abroad. We aimed to explore their experiences of the ongoing conflict. We wanted to understand the reasons for them to leave their home country after the conflict started in 2011.

### Study population

The qualitative study is based on semi-structured interviews (*n=20*) of Syrian HCWs who have left the country since 2011. The data was collected by three trained interviewers, one female and two males. Interviews (*n=17*) were conducted in Gaziantep, a Turkish municipality adjacent to the Syrian border where participants resided at the time of interviews in June – July 2016 and early 2017. Additional interviews (*n=3*) were conducted in Europe from July 2016 to early 2017. Altogether, participants consisted of 18 males and two females.

The age of participants ranged from 26 to 47 years. Mean age was 36 years. Most of the participants were born in Aleppo governorate (*n = 12*). The other participants represented a variety of other Syrian provinces. Most of the participants were married and had at least one child.

Majority of interviewed (*n = 14*) were physicians with post-graduate speciality (surgery and paediatrics were most typical) (*n = 9*). Also, other health care professionals, such as nurse (1), pharmacists (3) health service manager (1) and dentist (1) were interviewed. All interviewed had worked in Syria during the conflict at some point. The working experience as a health care professional, varied from almost none to over 20 years.

One of the participants was a medical student, and few general practitioners were residents in speciality training. They all had to suspend their studies because of the violence and leaving Syria.

### Study design and sampling

The participants were identified using a snowball sampling method (SSM), and they had to represent the category of health care workers in the International Labor Organization’s International Standards Classification of The Occupations (ISCO-08).[29] SSM was chosen to increase trust in the research that could be very sensitive.[30] Interviews were conducted in English or Arabic. Those undertaken in Arabic were later translated into English by a professional translator. The quality was confirmed by an independent translator using random extracts from the material. Each interview was terminated, when further inquiry provided no significant new themes.

### Measurement and analysis

A semi-structured interview guided the discussion with the participants. The study was focused on HCWs experiences of violence in Syria and reasons for migration from the country after 2011. The violence was defined in the study as “*the intentional use of physical force or power, threatened or actual, against oneself, another person, or against a group or community, that either results in or has a high likelihood of resulting in injury, death, psychological harm, maldevelopment or deprivation*.[31] We focused in this study health care workers’ experiences of violence. The attacks against health care were understood here as *an attack on health care as any act of verbal or physical violence or obstruction or threat of violence that interferes with the availability, access and delivery of curative and/or preventive health services during emergencies*.[32]

After the interviews were transcribed verbatim and triangulated with hand-written notes, the transcripts were read through. Using inductive content data analysis, answers were coded into categories-based experiences which indicated motivational factors for leaving the country. These factors were sought both from the direct answers to structured questions, but also from all of the free form replies. These were then classified into thematically unified sub-groups, no previously set system for such division was used, but the classification emerged organically from the population of factors introduced by the interviewees.

### Ethical approval

The interviews were recorded with permission from the participants, and consent was obtained verbally. Anonymity and confidentiality were bestowed. Participants were informed about the aim and purpose of the research. They were noticed that they are allowed to interrupt or abandon the interview if wanted. One of the interviewers was a psychotherapist. He monitored the mental state of the interviewees during the process.

An ethical permit for the study was applied for and obtained from the University of Eastern Finland Committee on Research Ethics. The principles in the Declaration of Helsinki were observed in all stages of the study.

### Patient and public involvement

No patients or members of the general public were involved in the conduct of this research.

## RESULTS

Reasons for leaving Syria were all related to violence that started after the demonstrations against the regime in 2011. The HCWs suffered either from general strain under the violent environment, be targeted by several specific violations or both. All participants expressed that their profession was the reason for targeting. The relative impact of these different forms of insults is further discussed in the conclusions part of the study.

The participants have gradually migrated during the conflict. Some left early when the violence started in 2011-2012, some later as the war escalated. While some moved to Eastern Aleppo, some moved directly to Turkey. Then many of those who remained initially in Eastern Aleppo followed. Some migrated to Lebanon due for geographical reasons. Once outside Syria, some HCWs continued to Europe to seek asylum.

### Generalised violence

When demonstrations escalated, and violence spread after 2011, the participants considered this as one of the turning points in their life. They described their experiences and feelings widely about what they had witnessed. They had seen atrocities against civilians, including their family and friends. People had been arbitrarily arrested and disappeared.

> *My friends were arrested in front of me. They were beaten, tortured in the streets. I saw these incidents. Everyone in Syria saw this. [The GoS was] killing people and shooting. Using tear gas, live bullets*. (general practitioner, male)

Before the territorial division between different warring factions, especially parting of NSAGs-controlled Eastern and GoS controlled Western side of Aleppo, had properly formed, the participants had regularly commuted through GoS checkpoints. The atmosphere had become oppressive. Constant checkups, arbitrary decisions, such as detaining, by the GoS and increasing threat of violence caused widespread psychological stress among the participants.

> *This fear of daily violence caused stress and anxious feelings. I could not sleep. I was very afraid. I decided to stop working. The reason for this was that I was stopped and bullied at the military checkpoint. I was asked ID card. All kinds of questions: “what you are doing in this city that is not your hometown and where does your family live”. They just wanted to bully me. Fear of being detained was the reason I left Syria*. (pharmacists, female)

A participant told that the situation became impossible for GoS to control. This resulting power vacuum gave NSAGs room to operate. Some participants expressed their mistrust of these organisations and couldn’t expect any help from the dwindling GoS force.

As the conflict evolved, participants witnessed barrel bombs, chemical attacks and airstrikes against civilians. They were not spared from these attacks.

> *I saw more than fifteen burnt Kurds, not far from the health care [centre]. Russian airstrike. Their relatives came. They didn’t [recognised them] because the bodies were burned. Disgusting*. (speciality doctor, male)

Other warring parties, mostly ISIS and NSAGs, conducted violence against civilians according to participants. While GoS was considered the main perpetrator, ISIS and other NSAGs, such as Free Syrian Army (FSA) and Jabhat al-Nusra threatened, mistreated, abducted and killed people.

> *FSA took one nurse, tortured him for five hours. Then he died. Just because his name was the same as some else’s*. (speciality doctor, male)

ISIS-inspired both fear and animosity against its ideology and values in the participants. The participant told unnerving anecdotes of the organisation that were circulating and creating an oppressive atmosphere of insecurity. Participants described their friends had severe problems with the organisation. Some of the participants decided to go to Turkey when they noticed ISIS advancing toward the city of Kobane, Aleppo governorate in northern east Syria. They believed fleeing from the organisation was their only option.

> *One of my friends was kidnapped [in Kobane], and the siege became more intensive. I knew that something is going to happen soon. When Kobane was besieged [by ISIS], I left for Turkey. I had to*. (nurse, male)

Some interviewees didn’t indicate a single reason for them leaving, but instead said they succumbed under the constant stress, fear and harassment because of the escalating situation.

### Violence related to personal issues

While participants were concerned about the generalised violence, their own and their families’ safety played an important role when they decided to leave Syria. When violence spread, many participants wanted to take their families into Turkey before fighting would reach their homes. Many noted that they wanted to keep their children safe, and in Syria, it was impossible.

> *I cannot live with my children under these circumstances. I saw what happened in Homs. I saw what happened in Deraa…using guns and aeroplanes… I felt this will happen in my area. This happened for 2-3 days [after leaving] my home*. (health service manager, male)

If a family member was arrested, all were investigated. Any suspected anti-government views or actions could lead to family-wide punishments, according to participants.

> *After the second time detention [by GoS] my family said: “Leave the country now”. I left. My family was afraid that I will go to jail again. Four days after I had left, they took my brother and told him “Tell your brother that if we catch him”…* (speciality doctor, male)

In many cases, the interviewed man was the breadwinner of the family. They were concerned about the financial well-being of their family, should they die. One participant mentioned that without a wife and children, he would most likely live and work in Syria.

> *Family is depended on me. Their existence is depending on me and my surviving. I don’t want to cause them danger or die as a martyr, and they lost me. It would be my family that would pay the high price*. (speciality doctor, male)

Some male participants expressed the fear of being conscripted in the Syrian forces. Many participants had witnessed atrocities performed by the GoS, and some had even been detained and tortured. They wanted to avoid becoming part of the military forces, and the only option was to leave.

> *We had to go. I knew that they [the GoS] were after me. I had to leave my studies and leave. I should have gone to the army. That’s why the regime was after me*. (speciality doctor, male)

Becoming remarked by ISIS for expressing opposing sentiments or antagonising them in any way was highly dangerous. Those who had fallen in their disfavour had little option. They had to leave because of being arrested or even executed.

> *We were talking about ISIS; that they are not from Syria and we are not accepting their presence here etc. Later my friends told me that ISIS is observing my home. I moved to another area. After this incidence, one of them [friends] was arrested by ISIS*. (specialist doctor, male)

Although the participants were not explicitly asked for their ethnic background because of the sensitivity, several them brought up their Kurdish roots, the minority in Syria. These Kurdish participants felt that the conflict had caused them to become targeted explicitly by radical Sunni Muslim NSAGs because of their background.

### Violence related to being a health care worker

Most of the participants had personally experienced violence that they considered to be connected with their profession. All thought that their profession made them as a target. Participants described their experiences widely since 2011. The violence included verbal assaults, beatings, detaining and torturing. They had been shot at or been in an ambulance when assaulted. Some described the situation when they had been in a health care facility in the time of the aerial bombardments by GoS, and later by Russia.

> *This [name retracted] hospital was targeted by a Russian airstrike. One of my friends died in this airstrike, the doctor [name retracted]. I’m very sad about him*. (speciality doctor, male)

All participants described the violence that their colleagues had experienced, and those were similar to their own. Some colleagues and co-workers were arrested, had gone missing and never seen later, some found dead with marks with severe abuse. Participants had also lost their colleagues in airstrikes.

> *I have seen colleagues killed in front of my eyes. In [place retracted] was a doctor, and she was taken. They [the unknown perpetrator] took her. Later she was found raped and dead*. (speciality doctor, male)

Participants saw and experienced their colleagues being humiliated and occasionally arrested in the hospitals, sometimes in the middle of medical operations.

> *[Name retracted] was like a brother. He got arrested in a real, humiliating way. He was changing the bandages to the patient. They [GoS] took him from the room. [They] covered his face in front of the staff of the hospital. They didn’t let anyone talk to him or ask where [GoS] were taking him*. (speciality doctor, male)

Some participants made their decision to leave once their colleagues had been arrested, gone missing or found dead. The fear of being caught by the GoS was a significant reason to escape. Many were afraid about their captured colleagues giving up their name under torture. However, a relationship marked by solidarity existed among the professionals.

> *One of my colleagues was arrested. Under a lot of pressure, they have their methods that you talk. He mentioned my name. I had to pay to get out of Syria and out of Aleppo. I left everything behind*. (general practitioner, male)

Also, as a HCW, participants were concerned the roadblocks. This was considered stressful when travelling for work. Participants described the incidences on the checkpoints. Sometimes they were stopped for more extended periods and inspections, sometimes detained. They considered that this was due to their profession.

> *At the [GoS] checkpoints, many doctors were arrested. One orthopaedic and his wife. This doctor had no problems [with GoS], and he had done nothing. He was taken from the roadblock. He was in prison for five months. His wife had to pay 800 000 Syrian pounds to get him away. This was in 2013*. (general practitioner, male)

Hospitals became threatening due to constant military and GoS presence. Armed soldiers were experienced as intimidating and reduced the willingness of HCWs to show up at work. One of the interviewees described having seen sharpshooters on hospital roofs aiming at people and even shooting them.

> *In the hospital where worked, the situation changed. It started to look like a military base. The soldiers were going in and out with their weapons. Most of the doctors and other health care workers did not come to work because they were afraid of the soldiers with guns. They could cause you troubles for no reason*. (speciality doctor, male)

In many cases, when the governmental forces arrived to investigate the hospital, HCWs warned their colleagues in danger of being arrested. The warning allowed them to avoid being captured. This was very dangerous, and in several cases, the interviewees reported GoS had arrested their colleagues.

> *I heard that my name was asked in the hospital I am working. I felt that I was in danger. I fled out of the country. When someone is asking about you in Syria, that is the mukhabarat [security service]. My friend was arrested in front of the hospital. One week before his arrest, they start to ask about him. One nurse in the hospital called me and [said] “There is someone from the military department and asked about you”*. (general practitioner, male)

The opposite was also true. A fraction of the workers sided with the GoS according to participants. They could spy on their colleagues, record their conversations and then turn them in. Due to strict internal monitoring anti-government discussions, even in private settings, could be carried to government officials.

> *They [security forces] opened the phone, and there were conversations between doctors. Some doctor or nurse have recorded conversations and then give them to the security service. We were arrested for 24-28 hours*. (speciality doctor, male)

While many left Syria under indirect threat, some of the professionals were persuaded to abandon their country only after having been personally threatened or imprisoned – some for extended periods.

> *After [released from the prison after six months], another intelligence department [officer] came to my house. They were looking for me. They were asking for my house -again. I left Aleppo to Turkey*. (speciality doctor, male)

A few years after the beginning of the conflict, other warring parties started causing problems for HCWs. ISIS was considered the most significant threat among the GoS.

> *ISIS arrested me. It was scary. In 2014 in Tel Abyad. For seven hours, then they let me leave. This was the main reason I decided to leave Syria. After being interrogated by ISIS, I decided that even I have studied for 12 years, I am not ready for this getting 1000 to 2000 USD per month [salary]. Those seven hours they interrogated me was a changing moment in my life. It was the first time when something like that happened to me. Those seven hours felt like seven years…* (speciality doctor, male)

HCWs are more likely to encounter events and victims of mistreatment due to the very nature of their profession. They witnessed the abuse of civilians while practising their profession. The interviewees describe having seen people subjected to violence and taken by GoS. Many of those persons imprisoned had been tortured before they were brought to hospitals.

> *We have seven patients taken by intelligence. We were receiving patients from the intelligence, after torturing in prison to cure them. Sometimes they bring dead bodies and throw them in front of us. Sometimes they took patients from the hospitals. Somebody who went to protest was shot or stabbed by Shabbiha [pro-government militia]. He [patient] came with his family or ambulance or by himself to the hospital to take treatment. They knew [GoS] that he was there, and they would come. Even before treating him. Or while treating him. I know many cases*. (speciality doctor, male)

## DISCUSSION

To our knowledge, this is the first research to examine the reasons why Syrian health care workers migrate from the conflict-affected country. This qualitative study presents the perspectives of the 20 Syrian HCWs who left the country at some point of the conflict. All participants left Syria because of security reasons. HCWs considered their profession as the reason for getting targeted. They migrated because of this, but also due to other personal issues and for the generalised violence.

While among the warring parties ISIS and NSAGs were mostly responsible for generalised violence and sometimes acted against the participants for personal reasons, the GoS seemed to target HCWs specifically because of their profession. This is in accordance with multiple reports and studies.[7,14,22,23,25] Many HCWs had to weigh the lack of prospects and accumulating stress against the equally intimidating challenge of actually trying to leave the country. While some participants had a specific notable trigger event, such as a colleague being detained or killed, many just grew tired of trying to manage under the constant threat and fear.

Financial issues and concerns regarding training were mentioned as reasons for leaving.[19] In this study, however, we found no HCWs indicating either factor as the primary cause for leaving. Doocy et al.[27] state that the choice to go is practically always a sum of many different factors. Our study suggested that all reasons were related to security issues. It appears the ever-present violence and complexity of the war in Syria supersedes all other concerns.

The average participant attending this study was a 36-year old male with a family and a medical doctor with a speciality. This profile is in accordance with other studies[20,21,27] that have shown male gender and similar age structure to associate with the risk of targeted violence and migration, and on the other hand, the will of protecting the family as an important reason to leave. As one of our participants said, he would be working in Syria if he did not have a family. When experienced HCWs leave the country, the remaining personnel are left without sufficient professional expertise as has happened in Iraq.[1] This reduces the quality of health care services and adds to the workload of those staying.[15,25]

The emotional distress of violence on HCWs is enormous. Participants have lost colleagues, friends and family members. In addition to their experienced traumas, they are most likely to have secondary traumas through witnessing atrocities against civilians while working as HCWs near the front-line of war. Despite this, some of our participants were visiting monthly in Syria as a humanitarian worker. The motivations for this should be studied more closely because it might help find solutions on how to get senior HCWs back permanently to Syria now, as the violence starts to show signs of decreasing. The availability of health care personnel is one of the central issues of rebuilding civil society. However, finding qualified HCWs is challenging when the majority of them have left the country.[15] Also, medical students had to leave their undergraduate studies and postgraduation specialisation training as they fled the violence.[17] These amount to a significant loss of human capital for the Syrian health care system. Restoring both current and future provision of services to an acceptable level will be extremely demanding.

## CONCLUSION

This research adds detailed information on how HCWs have experienced violence in conflict-torn Syria. This study gives a voice to Syrian health care workers who have witnessed horrors of conflict with extensive destruction and subsequently had no alternative but to leave their homes and work to protect themselves and their families. A better understanding of this type of forced migration is needed to develop approaches to support HCWs psychologically and practically in a time of war.

Our interviewees described such violence and attacks against health care that may constitute violations against Geneva Conventions. Thus, these actions might be considered war crimes and would then require perpetrators to be held accountable.

## Data Availability

All available data can be obtained from the corresponding author

## Strengths and limitations

The present research is subjected to several limitations. Firstly, while we reached participants from several different Syrian governorates (Homs, Daraa, Deir Ez-Zour, Raqqa, Hama, Rif Damascus), most of were originally from Aleppo. On the other hand, they had worked in different locations and had experienced violence in areas under all warring parties. More studies should be obtained from all these areas, especially from those that had been controlled by ISIS. Many interviewees considered ISIS as one of the main factors for leaving. Secondly, the snowball sampling method is conducive to selection bias.[30] We used three different parallel SSM networks to reduce this. Thirdly, the perspectives of female HCWs are partly missing. Future research should concentrate not only on gender-based violence but also on the experiences of female HCWs and their reasons for leaving Syria.

Obtaining data in a conflict-setting, especially first-hand accounts of personal experiences, is challenging. One of the authors is a Syrian HCW who has lived and worked mostly in NSAG and Kurdish forces-controlled areas. His first-hand knowledge made this study carry more weights this study and gives us a better understanding of the situation in such areas of Syria which researchers generally have not been able to visit due to the presence of severe security risks.

## ACKNOWLEDGEMENTS

The authors would like to sincerely thank all those brave health care workers who trusted us and shared their experiences with us.

## Contributors

AK, MH and JK designed and conceptualised the study. KA, MH and OA, HM coordinated and carried out data collection. AK, OA, and JP analysed and interpreted the data. AK led manuscript writing with contributions from OA, JP, MH, HJ and JK. All authors reviewed the final manuscript.

## Disclaimer

Foundation of the Finnish Institute in the Middle East was not involved in any step of this research. The notions and views are those of the authors.

## Patient consent

No patients or members of the public were involved in the conduct of this research

## Ethics approval

An ethical permit for the study was applied for and obtained from the University of Eastern Finland Committee on Research Ethics in 2016.

## Provenance and peer review

Not commissioned; externally peer-reviewed.

## Data sharing statement

All available data can be obtained from the corresponding author.

